# An Efficient and Interpretable Foundation Model for Retinal Image Analysis in Disease Diagnosis

**DOI:** 10.1101/2025.02.19.25322447

**Authors:** Wei Dai, Zhen Ji Chen, Yinghao Yao, Yu Chen, Jiyuan Fang, Qingshi Bai, Chuang Xu, Huimin Wu, Huaiyuan Ding, Hui Yang, Ran Zhuo, Riyan Zhang, Jian Yuan, Cong Ye, Hong Wang, Liangde Xu, Yongxin Yang, Xiaoguang Yu, Timothy Hospedales, Jia Qu, Jianzhong Su

**Affiliations:** State Key Laboratory of Eye Health, Eye Hospital, Wenzhou Medical University, Wenzhou, China; Oujiang Laboratory, Zhejiang Lab for Regenerative Medicine, Vision and Brain Health, Wenzhou, China; Department of Ophthalmology, The Second Affiliated Hospital and Yuying Children’s Hospital of Wenzhou Medical University, Wenzhou, China; Queen Mary University of London, UK; Institute of PSI Genomics, Wenzhou, China; School of Informatics, University of Edinburgh, UK

**Author notes:** These authors contributed equally (Wei Dai, Zhen Ji Chen, Yinghao Yao). Corresponding authors: Jia Qu, Jianzhong Su.

**Keywords:** Hierarchical representation learning, Contrastive learning, Clinical explainability, Resource-efficient

## Abstract

Artificial intelligence (AI) foundation models for colour fundus photography (CFP) have been extensively studied and demonstrated great potential for advancing ocular and systemic health screening. However, their high computational demands and limited clinical interpretability constrain real-world clinical application. These models rely on self-supervised learning with massive unlabeled datasets to address the scarcity of high-quality annotations, but often generate irrelevant features and fail to improve interpretability due to the absence of medical knowledge integration. Thus, we propose HRVRL, a lightweight, knowledge-prompt foundation model that leverages a novel hierarchical representation learning framework based on retinal biological features. Over 150,000 instances were generated for pretraining through multi-level image augmentation of 267 vascular-labeled images. A progressive learning strategy enables HRVRL to capture retinal-specific features from coarse to fine scales. HRVRL demonstrates remarkable resource efficiency, requiring only 0.04 GB of memory, processing 24 images per second, and completing pretraining within one day using a single GPU. It outperforms existing foundation models in 20 of 24 downstream tasks related to ocular and systemic disease diagnosis and severity grading. HRVRL also offers high clinical interpretability, with quantitative assessments showing strong concordance between model predictions and clinical criteria and outperforming in all 10 tasks. In diabetic retinopathy (DR) analysis, HRVRL achieves superior diagnostic lesion recognition (median accuracy of 0.710 versus 0.1–0.235 for existing models; *P* < 0.001) and significant improvements in type-specific lesion detection under a zero-shot setting (18-fold for hemorrhages, 4-fold for microaneurysms, hard exudates, and soft exudates; *P* < 0.001). We demonstrate that HRVRL provides clinically interpretable predictions with transparent decision-making processes for individual cases. In conclusion, HRVRL achieves unprecedented resource efficiency and enhanced clinical interpretability, enabling practical deployment in resource-limited settings to improve ocular and systemic disease diagnosis.

## Introduction

Color fundus photography (CFP) represents a cornerstone imaging modality in ophthalmologic practice, esteemed for its non-invasive nature, granular visualization of the retinal architecture, and robust diagnostic efficacy across a broad spectrum of ocular and systemic diseases^1,2^. Its seamless integration with telemedicine platforms, combined with minimal operational costs and standardized acquisition protocols, has transformed eye care accessibility, particularly facilitating early disease detection in resource-limited setting^3,4^. Recently, the advent of artificial intelligence (AI) foundation models, pretrained on extensive datasets to learn general representations that can be adapted to a variety of downstream applications, has instigated a transformative shift in the landscape of CFP analysis, significantly augmenting the detection capabilities of ocular and systemic conditions^5–10^. Notably, RETFound, a Vision Transformer (ViT)-based foundation model, has achieved remarkable diagnostic capabilities by learning generalized representations from 904,170 unlabeled real-world CFPs^11^. However, such unprecedented data requirements and high training cost pose significant barriers to developing custom foundation models in resource-constrained healthcare settings, while limited model interpretability presents fundamental obstacles to clinical deployment^11–15^.

Recent computational advances have markedly improved the efficiency of foundation models in retinal image analysis while preserving diagnostic performance. Through the integration of clinical domain knowledge for synthetic data generation, DERETFound reduced the requisite pretraining dataset to 150,786 raw images^16^. Subsequently, RETFound-Green further decreased the pretraining data requirement to 75,000 images through the implementation of an optimized token reconstruction method on a smaller Vision Transformer (ViT) architecture^17^. Despite these advances in data efficiency, current foundation models still face two fundamental limitations in CFP analysis that impede their clinical translation. The first limitation stems from their reliance on ViT-based architectures, which inherently require substantial pretraining datasets despite optimization efforts. The architectural constraint has motivated exploration of alternative approaches, such as ConvNeXt, which combines the inherent inductive biases of convolutional neural networks (CNNs) with Transformer-style global attention mechanisms. This hybrid architecture achieves comparable performance with significantly reduced computational demands, offering a more efficient solution^18^. The second limitation lies in the learning approach. Current models rely on generic self-supervised objectives, primarily masked pixel and feature reconstruction, without leveraging the rich anatomical patterns inherent in retinal images. This misalignment between computational objectives and domain-specific medical knowledge undermines both model interpretability and the extraction of clinically meaningful features from CFPs, constraining their practical value in healthcare settings.

In clinical practice, the incorporation of established medical knowledge, including pathological features and clinical presentations, forms the cornerstone of reliable diagnostic decision-making^19^. In CFP analysis, In CFP analysis, retinal vessels, fundamental anatomical structures of the retina, and their associated changes provide essential insights into both ocular and systemic diseases. Microaneurysms serve as a characteristic marker of diabetic retinopathy (DR), while vessel narrowing and arteriovenous nicking strongly correlate with hypertension and cardiovascular conditions^20,21^. These clinically validated patterns could serve as meaningful prompts for AI-driven models, potentially reducing training complexity while enhancing both accuracy and interpretability^22–24^. However, the effective integration of these domain-specific knowledge into current foundation models remains unexplored.

In this work, we propose an efficient and interpretable <u>H</u>ierarchical <u>R</u>etinal <u>V</u>ascular <u>R</u>epresentation <u>L</u>earning (HRVRL) foundation model for CFP analysis. HRVRL incorporates retinal-specific knowledge through a hierarchical learning framework using only 267 manually vascular-labeled images and a computationally efficient ConvNeXt backbone. The framework consisted of three key components: 1) multi-level data preprocessing to augment images; 2) a multi-level pretraining strategy that combines instance-level contrastive learning with pixel-level segmentation to capture arterial, venous, and non-vascular general latent representations (**Fig. 1a**) and 3) a multi-region interpretability method that quantifies and visualizes both regional and global contributions to model predictions (**Fig. 1d**). We adapted HRVRL to complex diagnostic tasks with explicit labels through two approaches: fine-tuning via transfer learning (fine-tuned) and feature extraction followed by a learnable multilayer perceptron (feature-based) (**Fig. 1b**). Comprehensive evaluations against competing CFP foundation models including RETFound, DERETFound and RETFound-Green in terms of training cost and downstream tasks performance (**Fig. 1c**) demonstrated that HRVRL achieves outstanding performance using the smallest pretraining dataset, the shortest pretraining time, minimal memory usage, and the highest inference speed. Notably, HRVRL achieves superior clinical interpretability that aligns closely with clinicians’ diagnostic patterns, enabling multi-type lesion detection in a zero-shot manner, potentially enabling advancements in ophthalmic and systemic health research.

**Fig. 1.**
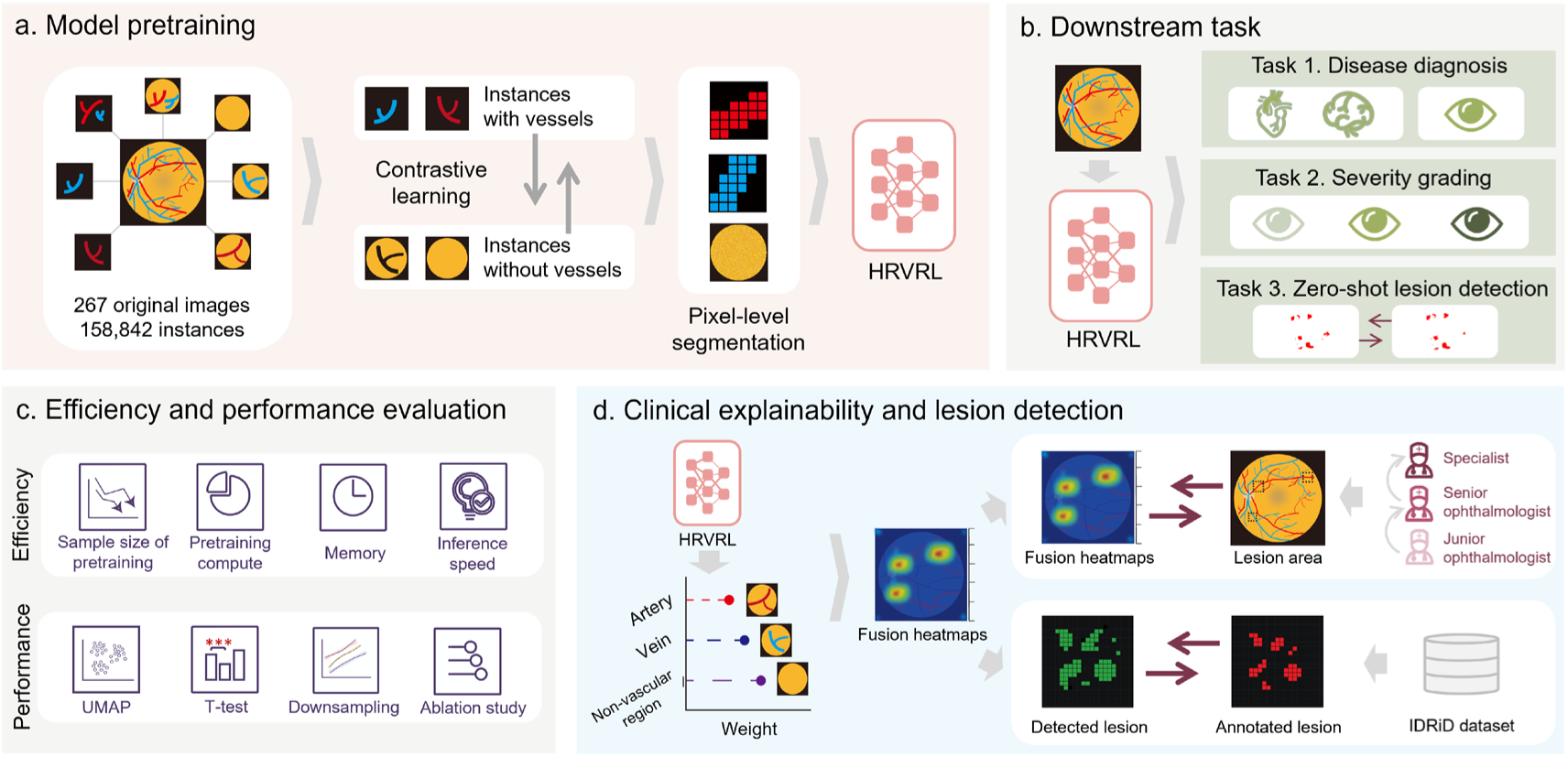
Overview of the study. **a**, Model pretraining. The hierarchical retinal vascular representation learning (HRVRL) model, built on a ConvNeXt backbone, employs a novel preprocess strategy, augmenting 267 original images to 158,842 instances, and a hierarchical pretraining strategy: an instance-level learning by contrasting instances with and without vessels and then a pixel-level learning by segmenting each pixel into arterial, venous, or non-vascular regions. **b**, Downstream tasks. HRVRL is adapted for ocular and systemic disease diagnosis, multi-severity grading of ocular diseases, and performs lesion detection in a zero-shot manner. **c**, Efficiency and performance evaluation. We conducted efficiency comparisons between HRVRL and three data-driven models in terms of resource consumption and inference speed. The performance of HRVRL in pretraining and disease diagnosis was assessed using UMAP visualization, t-tests, downsampling analysis, and ablation studies. **d**, Clinical explainability and lesion detection. A hierarchical explainable method was proposed to quantitative and qualitive analysis of the contributions of artery, vein and non-vascular regions to model decision. A fusion heatmap was generated to visualize the clinical explainability of the models and to evaluate the similarity with lesion areas identified by retinal ophthalmologists. We further explored the ability of HRVRL to detect type-specific lesions on the public lesion segmentation dataset IDRiD without task-specific training.

## Results

### Overview of HRVRL hierarchical retinal vascular representation learning model

Figure 1 illustrates the comprehensive flowchart of HRVRL. The model was pretrained using 267 CFPs collected from diverse imaging devices and populations, with expert-labeled arterial and venous annotations (**Supplementary Table 1 and Methods**). The pretraining process of HRVRL involved two main steps (Fig. 1a). First, to enhance data efficiency, we devised a multi-level augmentation strategy, increasing the 267 raw images to 1,869 generated images, and subsequently into 158,854 representative instances across multiple scales, prompted by vascular knowledge (**Extended Data Fig. 1, 2 and Methods)**. Second, we employed instance-level contrastive learning to distinguish vascular from non-vascular features, followed by pixel-level semantic segmentation to differentiate arterial from venous features (**Extended Data Fig. 3 and Methods**). HRVRL was evaluated on three major downstream tasks: ocular disease diagnosis, systemic disease diagnosis, severity grading of retinal diseases, along with zero-shot lesion detection, using 12 independent datasets (**Fig. 1b, Supplementary Table 2 and Methods**). Furthermore, the efficiency, performance, and interpretability of HRVRL were benchmarked against three foundation models: RETFound, DERETFound, and RETFound-Green (**Fig. 1c**). For disease diagnosis and severity grading tasks, we evaluated both fine-tuning and feature-based approaches in adapting the pretrained foundation model. Finally, we analyzed the contributions of arterial, venous, and non-vascular regions to HRVRL’s decision-making through multi-regional heatmaps (**Fig. 1d and Methods)**. These heatmaps demonstrated consistency with clinical standards and effectiveness in zero-shot lesion detection. Complete benchmarking results are summarized in **Table 1**.

**Table 1.** Overall efficiency and performance comparisons among the HRVRL model and the state-of-the-art (SOTA) models.

|  | RETFound | DERETFound | RETFound-Green | HRVRL (Ours) | HRVRL vs SOTA |
| --- | --- | --- | --- | --- | --- |
| **Efficiency** |  |  |  |  |  |
| **Pretraining data** | 904,170 images | 150,786 images<br>(~1000,000 generated images) | 75,000 images | 267 images<br>(158,842 instances) | 280× less |
| **Pretraining compute** | 112 days (A100) | 163 days (A100) | < 1 day (A100) | < 1 day (A100) | Comparable |
| **Memory** | 1.12 GB | 1.12 GB | 0.09 GB | 0.04 GB | 2× less |
| **Input size** | 224 × 224 | 224 × 224 | 392 × 392 | 512 × 512 | 1.7× richer |
| **Inference speed** | 6 img/s | 6 img/s | 16 img/s | 24 img/s | 1.5× faster |
| **Performance** |  |  |  |  |  |
| **Classification (fine-tuned)**<br>(Win at $P < 0.05$, Fig. 2) | 0 win | 2 wins | 1 win | 9 wins | 4.5× the wins |
| **Classification (feature-based)**<br>(Win at $P < 0.05$, Extended Data Fig. 5) | 0 win | 1 win | 0 win | 11 wins | 11× the wins |
| **Clinical explanation**<br>(Win at $P < 0.05$, Fig. 4) | 0 win | 0 win | 0 win | 5 wins | Completely win |
| **Zero-shot lesion detection**<br>(Win at $P < 0.05$, Fig. 5) | 0 win | 0 win | 0 win | 5 wins | Completely win |

**Abbreviations:** A100, NVIDIA A100 GPU; GB, Gigabytes; img/s, image(s) per second.

### Model efficiency evaluation

To evaluate the efficiency of HRVRL against competing CFP foundation models, we analyzed pretraining data requirements, pretraining time, memory usage, and inference speed. HRVRL demonstrated significant advantages across all metrics (**Table 1**). First, it exhibited impressive data efficiency, requiring only 267 CFPs for pretraining, which was 280-fold less than RETFound-Green and over 3,300-fold less than RETFound. Second, HRVRL completed training from scratch with randomly initialized weights in less than one day on a single NVIDIA A100 GPU, dramatically faster than RETFound (112 days) and DERETFound (163 days) on the same hardware, and comparable to RETFound-Green despite its reliance on pre-initialized weights^17^. HRVRL also demonstrated superior memory efficiency, requiring only 0.04 GB of memory during training, half that of RETFound-Green and 22-fold less than RETFound and DERETFound. This reduced memory requirement can be attributed to the fewer learnable model parameters, which decreased computational overhead and further enhanced inference efficiency. HRVRL processed 24 images per second, outperforming RETFound-Green by 1.5-fold and RETFound by 4-fold in inference speed. Additionally, HRVRL operated at an increased input resolution of 512 × 512 pixels, providing enhanced spatial detail for downstream tasks. These comprehensive efficiency gains, combined with minimal data requirements and faster inference speeds, make HRVRL a scalable and practical solution for clinical deployment.

### Task-specific HRVRL performance evaluation

To assess HRVRL’s performance across various downstream tasks, we conducted both fine-tuning and feature-based analyses on 12 datasets: four multi-category ocular disease datasets, four binary systemic disease datasets, and four severity grading datasets. HRVRL demonstrated superior performance in 9 of 12 fine-tuning tasks and 11 of 12 feature-based tasks, highlighting both its strong adaptability and the quality of its pretrained representations (**Table 1, Fig. 2, Extended Data Fig. 4**). For multi-category ocular disease diagnosis, HRVRL was evaluated on PAPILA, Retina, BRSET, and UKBB datasets. We found that HRVRL (fine-tuned) outperformed other models on three of these datasets and ranked second on BRSET (all *P* ≤ 0.001; **Fig. 2a**), while HRVRL (feature-based) achieved the highest performance across all four datasets (all *P* ≤ 0.001; **Extended Data Fig. 4a**). In systemic disease detection, including heart failure, myocardial infarction, cerebrovascular diseases, and type 2 diabetes, all derived from the UK Biobank, both fine-tuned HRVRL and feature-based HRVRL achieved the highest performance, with RETFound and DERETFound ranking second (all *P* < 0.001; **Fig. 2b, Extended Data Fig. 4b**). For severity grading of diabetic retinopathy (DR), evaluated on Kaggle APTOS-2019, IDRiD, and MESSIDOR-2 datasets (severity grades according to five-stage International Clinical DR Severity Scale^21^), HRVRL achieved the best performance on two datasets and ranked second on one in fine-tuning evaluation (**Fig. 2c**) while outperforming all models in feature-based evaluation (**Extended Data Fig. 4c**). In glaucoma severity grading, HRVRL ranked second on the Glaucoma Fundus dataset. All details are shown in Supplementary Table 3 and 4, for example, fine-tuned HRVRL achieved an AUROC of 0.967 (95% CI: 0.960–0.973) in MESSIDOR-2, significantly outperforming RETFound (*P <* 0.001), while feature-based HRVRL achieved an AUROC of 0.958 (95% CI: 0.953–0.964) in Kaggle APTOS-2019. Besides, label efficiency evaluation revealed that HRVRL maintained superior performance with reduced training data. It achieved results comparable to the competing models on the heart failure and IDRiD datasets while using 90% and 85% of the training data, respectively. On MESSIDOR-2, HRVRL outperformed all competing models using only 65% of the training data (**Extended Data Fig. 5, Supplementary Tables 5**). These comprehensive evaluations demonstrate HRVRL’s exceptional performance and data efficiency across diverse clinical tasks, highlighting its potential for practical clinical applications.

**Fig. 2.**
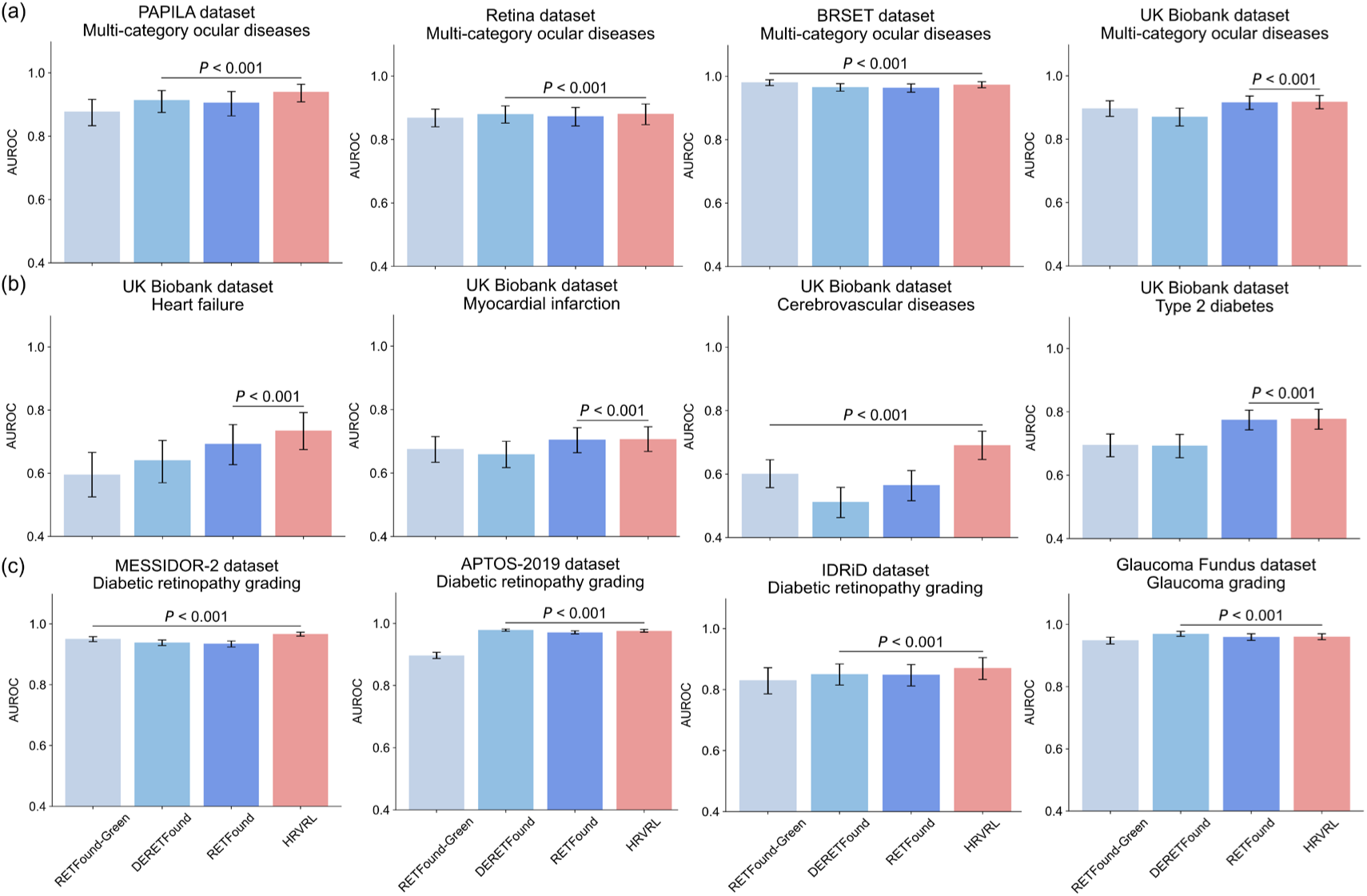
Performance on disease diagnosis and severity grading based on fine-tuning approach. **a**. Ocular disease diagnosis. The models are fine-tuned on four datasets (PAPILA, Retina, BRSET, and UK biobank datasets) separately to differentiate between healthy controls and various ocular diseases, such as age-related macular degeneration, diabetic retinopathy, glaucoma and hypertensive retinopathy. **b**. Systemic disease diagnosis. The models are fine-tuned to diagnose systemic diseases from the UK biobank dataset, including heart failure, myocardial infarction, cerebrovascular diseases and type 2 diabetes. **c**. Severity grading of retinal diseases. The models are fine-tuned to rank different severity of diabetic retinopathy (MESSIDOR-2, APTOS-2019, and IDRiD dataset) and glaucoma (Glaucoma Fundus dataset). These diseases categories and datasets characteristics are summarized in Supplementary Table 2. HRVRL outperforms other models in nine out of twelve downstream tasks and is ranked second in the remaining three. Error bars denote the 95% confidence intervals (CIs) surrounding the estimates, with the bar centers representing the mean values of the metric under consideration. These estimates were derived from a bootstrap distribution involving 1,000 times resamples of test dataset to ensure statistical significance. We compared the performance of HRVRL with the most competitive comparison model among RETFound, DERETFound and RETFound-Green to determine the presence of statistically significant differences. A two-sided t-test was conducted to evaluate the statistical significance of the differences observed.

### Hierarchical interpretation for task-specific HRVRL

To enhance the interpretability and clinical relevance of HRVRL’s predictions, we developed a hierarchical interpretation approach that integrated gradient-based saliency maps with Shapley values to quantify and qualify the contributions of arterial, venous, and non-vascular regions to model decision-making, respectively. Four exemplary diseases including two ocular diseases of hypertensive retinopathy (HR, N = 50) and DR (N = 30), and two systemic diseases of heart failure (HF, N = 60) and myocardial infarction (MI, N = 159) were involved in to evaluate the approach (**Supplementary Table 6** and **Methods**). For ocular diseases, our analyses demonstrated that HRVRL’s attention areas aligned with established diagnostic criteria (**Fig. 3**). In HR cases, fusion heatmaps revealed significant contributions from both vascular and non-vascular regions, with regional heatmaps showing venous curvature abnormalities as the dominant vascular features, while optic cup and disc were most significant in non-vascular regions (**Fig. 3a, Supplementary Table 6**). Shapley analysis revealed comparable regional contributions from venous (0.344), non-vascular (0.338), and arterial regions (0.297) (**Fig. 3b, Supplementary Table 6**), aligning with known HR biomarkers, such as arteriovenous nicking or nipping and microaneurysms^23^. In DR cases, non-vascular regions showed the highest contribution (0.439). It increased from the early (0.415) to the advanced stage (0.538), reflecting lesion progression in the respective heatmaps (**Fig. 3, Supplementary Table 6**). Additionally, venous contributions were lower than arterial in early stages but surpassed them in severe stages, consistent with venous beading appeared in advanced DR^21^. For systemic diseases, we observed that the contributions of arterial, venous, and non-vascular regions for HF were nearly equivalent (0.333, 0.336, 0.338, respectively, **Supplementary Table 6**). Fusion and regional heatmaps demonstrated that the model concentrated on both the optic cup and disc regions, as well as the adjacent vascular structures, suggesting that retinal vascular abnormalities and changes in the optic nerve head are crucial for HF diagnosis (**Extended Data Fig. 6**). In MI cases, vascular regions, both arteries and veins, dominated the model’s diagnostic contributions, with median Shapley values of 0.365 and 0.339, respectively, while non-vascular regions contributed less (0.288) (**Supplementary Table 6**). Heatmaps indicated a focus on vascular structures, such as vessel narrowing, is critical for MI diagnosis. These findings indicate that hierarchical interpretation approach of HRVRL effectively captures clinically relevant features, highlighting its ability to capture disease-specific retinal markers.

**Fig. 3.**
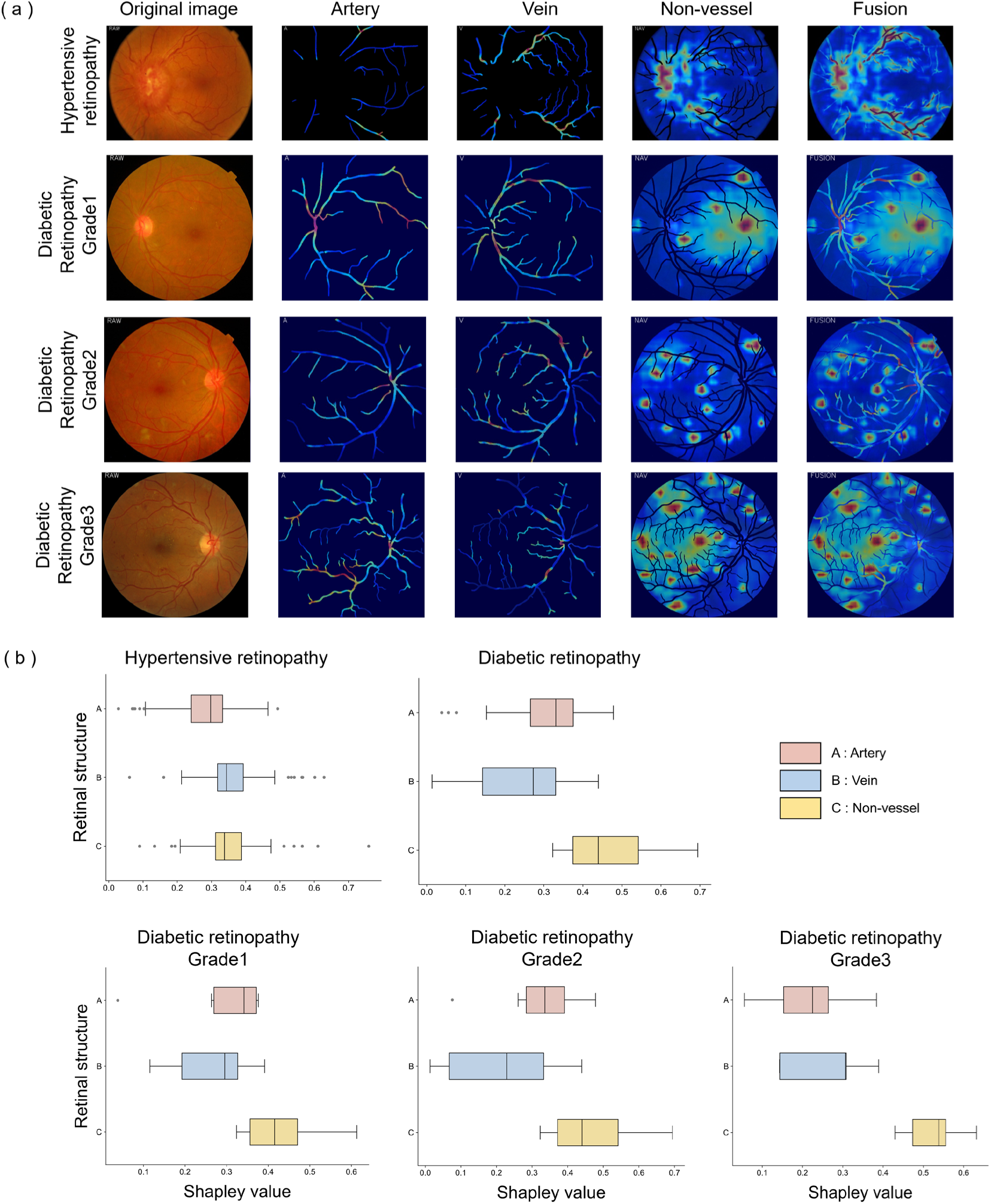
Results of qualitative and quantitative analysis for the contribution of different regions to the decision-making of fine-tuned HRVRL on exemplary ocular diseases. **a.** Qualitative regional heatmaps and fusion heatmaps generated by proposed hierarchical explainable method. The top row shows a case of hypertensive retinopathy (HR), and the following three rows illustrate different grades of diabetic retinopathy (DR). Each example from left to right depicts retinal images along with regional (arterial, venous, and non-vascular) heatmaps and fused heatmap. **b.** Quantitative contribution of retinal structures (artery, vein, and non-vascular region) to disease diagnosis. Box plots represent the overall Shapley values of each retinal region on HR and DR diagnosis across different severity grades, indicating the relative contribution of each region to HRVRL decision-making.

### Evaluation of consistency between HRVRL interpretations and clinical standards

To evaluate the consistency of HRVRL’s clinical interpretations with established standards, we compared model-generated heatmaps with expert annotations from four independent ophthalmologists on 30 HR and 30 DR images across varying severity grades^21,23^(**Methods**). The comparison focused on lesion detection rates between model predictions and expert annotations. **Fig. 4** showed the heatmaps for HR and DR generated by different models (top two rows in **Fig. 4a**), and the lesion detection rates for HR, overall DR, and multi-severity of DR (**Fig. 4b**). For HR, HRVRL precisely key diagnostic features including arteriovenous nicking, venous tortuosity, and arterial hardening, while other models were either distracted by irrelevant regions (RETFound and DERETFound) or showed limited focus (RETFound-Green). For DR, HRVRL accurately highlighted characteristic lesions such as microaneurysms and exudates, whereas other models either emphasized peripheral regions (RETFound), or struggled with noise and incomplete coverage (DERETFound and RETFound-Green). Quantitative analysis demonstrated HRVRL’s superior interpretability, achieving similarity scores above 0.65 with clinician annotations across diseases (all *P* < 0.001) (**Supplementary Table 7**). For HR diagnosis, HRVRL’s score of 0.667 was over six-fold higher than RETFound-Green (0.118), while in DR detection, HRVRL maintained consistent performance across severity levels, achieving a 0.75 lesion detection rate in severe DR cases, substantially outperforming DERETFound (0.472).

**Fig. 4.**
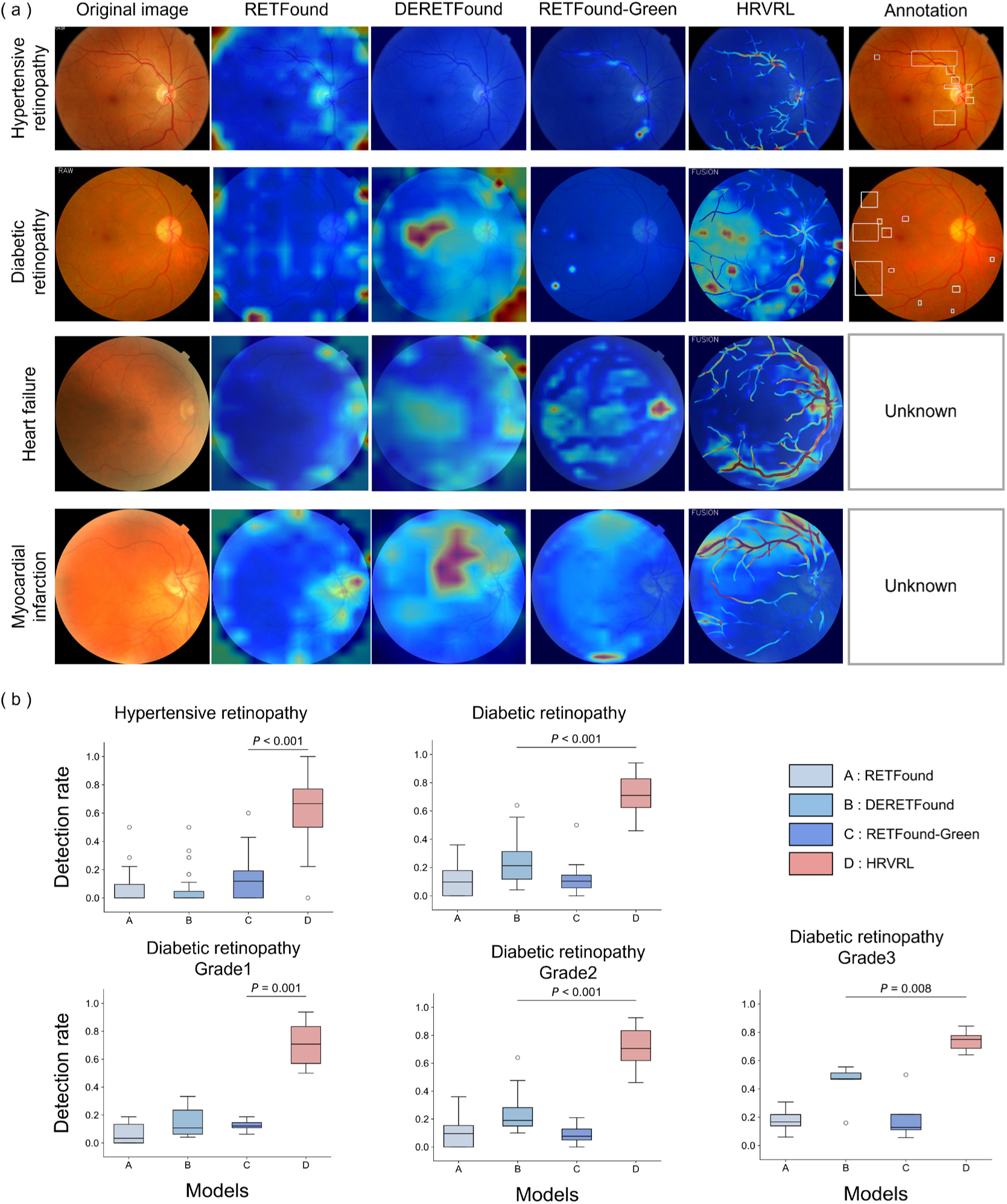
Performance assessment and comparison of clinical interpretability. **a.** The comparison of heatmaps about HRVRL, three foundation models: RETFound, DERETFound, and RETFound-Green with the expert annotation. Red area in heatmap indicates high contribution while blue means low contribution. Labels with white boxes, annotated by retinal specialists, are the markable symptoms of disease diagnosis. The top two row shows cases of ocular diseases HR and DR with clinical annotations. The bottle rows show cases of systemic diseases HF and MI without clinical annotations. **b.** The comparative analysis of different models in lesion detection rate. Box plots display the lesion detection rates for HR, DR (Grade 1, Grade 2, Grade 3) across RETFound, DERETFound, RETFound-Green, and HRVRL. A Mann–Whitney U-test was conducted to evaluate the statistical significance of the observed differences.

To further explore the HRVRL’s clinical explanations in systemic diseases lacking established criteria, we visualize the key areas influencing decision-making of models. In HF cases, HRVRL highlighted vascular alterations and optic area changes, aligning with known cardiovascular manifestations^20,23^. In contrast, heatmaps from other models primarily focused on the optic disk (RETFound-Green) or the image edges without focusing vascular information (RETFound and DERETFound) (third row in **Fig. 4a**). For MI, the heatmap of HRVRL showed a strong focus on arterial and venous alterations, particularly highlighting areas of vascular narrowing and high tortuosity (last row in **Fig. 4a**). In comparison, competing models concentrated mainly on the optic disc or non-vascular changes, which were less relevant to the alterations associated with MI^25^. These comprehensive evaluations demonstrate HRVRL’s consistent ability to identify clinically relevant features across both ocular and systemic diseases.

### Zero-shot detection of type-specific lesions

To assess HRVRL’s ability in detecting type-specific lesions, we conducted zero-shot analysis on 72 images from the IDRiD dataset with pixel-level lesion annotations (**Methods**). Using recall rate to measure overlap between model interpretation and expert annotations, HRVRL significantly outperformed other models in detecting key DR pathological features including microaneurysms, hard exudates, soft exudates and hemorrhages (**Fig. 5** and **Supplementary Table 8**). Overall, HRVRL achieved a recall rate of 0.518, substantially higher than DERETFound (0.134), RETFound (0.030), and RETFound-Green (0.004). For microaneurysms, the earliest and most challenging lesion in DR progression, HRVRL achieved a recall of 0.404, at least 4-fold higher than other models (*P <* 0.001). Similar superiority was demonstrated in detecting soft exudates (0.466) and hard exudates (0.555), both exceeding other models by more than 4-fold (*P* < 0.001). Most notably, in detecting hemorrhages, the most vision-threatening retinal alteration, HRVRL achieved an 18-fold improvement in recall compared to other methods (*P* < 0.001). In summary, HRVRL demonstrated superior performance in type-specific lesions detection, underscoring its significant potential for accurate disease detection aligned with clinical criteria.

**Fig. 5.**
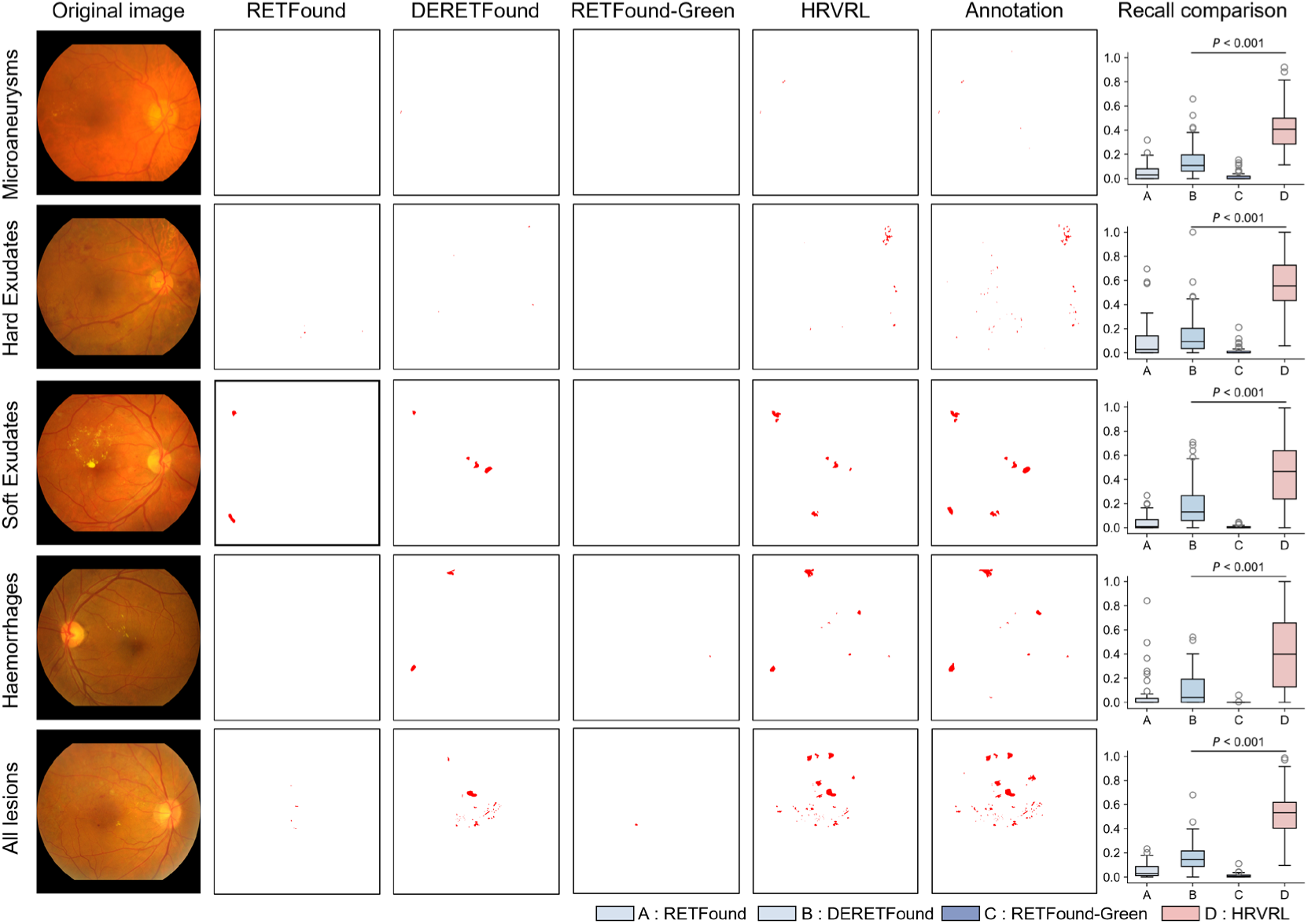
Performance assessment and comparison of zero-shot type-specific lesions detection. The figure provides a comprehensive comparison of different models for detecting various DR lesions without task-specific training. The first five columns show the original fundus images, followed by the lesion detection binary masks generated by RETFound, DERETFound, RETFound-Green, and HRVRL, where highlighted areas indicate the identified potential lesions. The sixth column displays the ground-truth annotations made by experts, which serve as the reference standard for comparison. Each row represents a specific type of lesion: microaneurysms, hard exudates, soft exudates, haemorrhages, as well as a summary row for all lesions combined. The rightmost column presents box plots comparing the lesion detection performance of these models across different lesion types. A Mann–Whitney U-test was performed to assess the statistical significance of the observed differences.

### Unveiling the mechanisms of high interpretability and performance of HRVRL

To understand the factors behind HRVRL’s superior performance and clinical interpretability, we investigated both the learned latent representations during hierarchical pretraining and the impact of different pretrained weights on downstream tasks. Visualization analysis revealed that HRVRL effectively acquired retinal-specific knowledge during pretraining, successfully differentiating between vascular and non-vascular features during instance-level contrastive learning. The model’s ability to distinguish arterial, venous, and non-vascular structures was further demonstrated through semantic segmentation visualizations on both internal and external datasets (**Extended Data Fig. 7**). Comparative analysis of different pretraining strategies, including random initialization, supervised learning on ImageNet-1k (SL-ImageNet), and instance-level contrastive learning of HRVRL (ICL-HRVRL), highlighted the advantages of domain-specific knowledge-guided pretraining (**Extended Data Fig. 8**). For example, ICL-HRVRL, which integrated intermediate levels of medical knowledge during pretraining, achieved an AUROC of 0.86 in DR classification (IDRiD dataset), compared to 0.80 with SL-ImageNet (**Supplementary Table 9**). This improvement underscored that even partial incorporation of domain-specific retinal features, such as vascular structures, leads to enhanced performance. HRVRL further improved this to 0.91, demonstrating that embedding comprehensive domain-specific retinal features, such as vascular structures, enhances the performance of downstream task. These findings underscore the crucial role of medical knowledge integration in pretraining for achieving superior task-specific performance and clinical applicability.

## Discussion

The development of current CFP foundation models has been limited by their substantial computational demands and lack of clinical interpretability. Here, we present HRVRL, which addresses these challenges through a novel knowledge-guided pretraining approach. By incorporating domain-specific vascular knowledge within a hierarchical learning framework, HRVRL achieves superior performance using just 267 vascular-labeled images, demonstrating that targeted knowledge integration can effectively overcome the data-hungry nature of current foundation models. Specifically, HRVRL is the best model in 30 out of 34 downstream tasks and ranked second in the remaining four (**Table 1**). Besides, HRVRL aslo provides both regional and global explanations for its predictions, enhancing transparency and aligning closely with clinical insights. Our findings demonstrate the potential of HRVRL in the clinical diagnosis of ophthalmic diseases and systemic diseases and offer new methodologies of knowledge-guided hierarchical pretraining that can be applied to other areas of medical imaging, advancing the field of medical artificial intelligence, especially in resource-constrained settings.

HRVRL fundamentally differs from existing CFP foundation models in both methodology and efficiency. In detail, current foundation models rely on masked pixels or features reconstruction methods as pretraining tasks, which assign equal importance to all pixels and features. This indiscriminate approach not only requires large amounts of pretraining data but also introduces substantial noisy or redundant information, significantly increasing the size of model parameters and computational demands. In contrast, HRVRL takes a fundamentally different approach by employing a hierarchical pretraining strategy that progressively learns vascular features from global to fine scales (**Extended Data Fig. 7**). This knowledge-driven framework focuses on capturing essential retinal structures while avoiding noise accumulation, thereby addressing the inefficiencies of traditional models. As a result, HRVRL reduces the size of model parameters to just 0.04 GB, which is significantly smaller than competing models, and achieves pretraining from scratch on a single GPU within a single day. This shift toward knowledge-guided, resource-efficient pretraining highlights the potential of such approaches to democratize AI development in medical imaging.

HRVRL’s clinical value extends beyond its computational efficiency (**Fig. 3–5 and Extended Data Fig. 6**). In ocular diseases like DR, where severity grading relies on well-defined criteria including lesion types, distribution, and counts^21^, HRVRL demonstrates unprecedented alignment with clinical criteria, significantly outperforming existing models in detecting specific pathological features. We attribute this improved consistency largely to the incorporation of medical knowledge during the pretraining phase. This consistency with clinical guidelines ensures its outputs are actionable in real-world clinical workflows, significantly enhancing the trust and practicality of AI-based diagnostic tools. For systemic diseases such as HF and MI, where established clinical imaging standards are often lacking, HRVRL demonstrates its capability by identifying key vascular features associated with systemic conditions^26–30^. This capability allows clinicians to gain deeper insights into disease mechanisms and showcases HRVRL’s potential as a tool for early disease detection and monitoring. By dynamically focusing on relevant features, HRVRL not only predicts disease outcomes but also offers valuable insights into systemic disease progression. This makes it a powerful framework for investigating clinical scenarios where decision-making lacks clear, well-defined guidelines.

However, several limitations of our current work suggest directions for future development. First, while HRVRL effectively leverages vascular annotations, incorporating more detailed retinal structure annotations could further enhance its capabilities, particularly for specific pathological assessments. Second, while HRVRL uses vascular masking to emphasize non-vascular regions, vascular features like diameter and curvature still affect predictions. Future efforts should refine masking strategies to better isolate non-vascular features, enhancing clarity in pathological processes and improving prediction interpretability. Third, HRVRL is designed for CFP analysis but lacks integration with other modalities such as optical coherence tomography (OCT), optical coherence tomography angiography (OCTA), fundus fluorescein angiography (FFA), and clinical reports. Incorporating multimodal data could provide more detailed information. Future approaches that combine CFPs with other modalities may enhance HRVRL’s ability to comprehensively analyze diverse retinal pathologies.

In conclusion, the success of HRVRL in achieving high performance with minimal data and computational resources while maintaining clinical interpretability has broader implications for medical AI development. Our results demonstrate that incorporating domain-specific knowledge can effectively address the limitations of data-hungry foundation models, potentially extending this approach to other medical imaging domains where large datasets are unavailable or impractical to obtain. This work marks a significant advance for developing resource-efficient, clinically aligned AI systems, particularly valuable for healthcare settings with limited computational resources.

## Methods

### Datasets for developing HRVRL

We employ a small collection of color fundus photos (CFPs) (267 images) with arterial and venous labels by experts for the HRVRL model, which are from four public datasets (207 images, 77.53%) and one in-house dataset (60 images, 22.47%) to enhance the generalization of the HRVRL across different devices (**Supplementary Table 1**). The public datasets comprise 40 images (14.98%) from AV-DRIVE^31^, 22 images (8.24%) from LES-AV^32^, 45 images (16.85%) from HRF^33^, and 100 images (37.45%) randomly selected from UK Biobank^34^ (UKBB). The in-house dataset comprises 60 randomly selected images from the Myopia Associated Genetics and Intervention Consortium (MAGIC) cohort (22.47%)^35^. The fields of view (FOVs) of retinal imaging vary (**Extended Data Fig. 2**), such as 30-degree FOV (21 images, 7.87%), 45-degree FOV (171 images, 64.04%), 50-degree FOV (30 images, 11.24%) and 60-degree (45 images, 16.85%). The imaging devices are diverse, including CR5 nonmydriatic 3CCD camera (CANON), CF–60 UVi (CANON), Visucam ProNm fundus camera, 3DOCT-1000-Mk2 (TOPCON) and RetiCam 3100 (**Supplementary Table 1**). AV-DRIVE, LES-AV and HRF are public datasets for developing algorisms of artery and vein segmentation models; thus, they have expertized vascular labels. For the UKBB and MAGIC datasets, four ophthalmologists labeled the retinal arteries and veins.

### Data for ocular and systemic disease diagnosis

We evaluate the model performance of the HRVRL on various tasks related to disease detection. The first task involves the diagnostic classification of both ocular and systemic diseases using publicly available datasets. we utilized several publicly accessible datasets encompassing multiple diseases, including UKBB (United Kingdom), Retina, PAPILA (Spain), and BRSET (Brazil). UKBB is a population-based cohort of over 500,000 participants recruited from 2006 to 2010, each undergoing retinal fundus imaging at enrollment. Disease diagnosis within the UKBB is based on the 1,866 hierarchical phenotypes defined by the Phecode Map 1.2 ICD-9 (https://phewascatalog.org/phecodes) and ICD-10 (https://phewascatalog.org/phecodes_icd10) phenotype grouping^36,37^. From the UKBB, we selected participants diagnosed with one of the following four ocular diseases: age-related macular degeneration (AMD, phecode: 362.29), diabetic retinopathy (DR, phecode: 250.7), glaucoma (phecode: 365), and retinal vascular occlusion (RVO, phecode: 362.4), while controls were randomly selected from a previous study and defined as having a healthy ocular condition^38^. The Retina dataset includes labels for normal eyes, glaucoma, cataract, and other retinal diseases. PAPILA contains labels for glaucoma, suspected glaucoma, and normal eyes, as determined by two experts following extensive clinical examinations^39^. From BRSET, we select cases of AMD, hypertensive retinopathy (HR), diabetic macular edema (DME) and healthy controls without ocular or systemic disease labels^40^. For systemic disease classification, we focused on six types of diseases known to be highly correlated with retinal changes^4,41,42^: type 2 diabetes (T2D, phecode: 250.2), hypertension (HT, phecode: 401), heart failure (HF, phecode: 428.1 and 428.2), myocardial infarction (MI, phecode: 411.2), Parkinson’s disease (PD, phecode: 332), and cerebrovascular disease (CeVD, phecode: 430 and 433). Retinal images are from UKBB dataset, diagnosed with type 2 diabetes (T2D, phecode: 250.2), hypertension (HT, phecode: 401), heart failure (HF, phecode: 428.1 and 428.2), myocardial infarction (MI, phecode: 411.2), Parkinson’s disease (PD, phecode: 332), and cerebrovascular disease (CeVD, phecode: 430 and 433). Retinal images from the UKBB dataset were used, and based on the sample sizes for each diagnosis, images were randomly selected using a case-control ratio of 1:1. Controls were randomly chosen from a previous study and defined as having a healthy ocular condition^38^. The detailed characteristics of used ocular and systemic diseases are shown in **Supplementary Table 2**, including imaging devices, country, label category and sample size.

### Data for severity grading of ocular disease

For the second task of ocular disease grading, datasets with severity ranks of diabetic retinopathy (DR) and glaucoma are involved in. We apply IDRiD (India), MESSIDOR-2 (France) and APTOS-2019 (India, Kaggle dataset) to evaluate the performance of DR grading^43,44^. The labels for DR are based on the International Clinical Diabetic Retinopathy Severity scale, indicating five stages from no diabetic retinopathy to proliferative diabetic retinopathy. For glaucoma, Glaucoma Fundus (South Korea) are included. Glaucoma Fundus has three categorical labels, non-glaucoma, early glaucoma and advanced glaucoma^45^. The grading protocols for the public datasets are summarized as: IDRiD, two medical experts provided adjudicated consensus grades; MESSIDOR-2, adjudicated by a panel of three retina specialists in accordance with a published protocol; APTOS-2019, Kaggle dataset with limited information but possibly a single clinician grader; Glaucoma Fundus, agreement of two specialists based on visual fields. The characteristics of the datasets are summarized in Supplementary Table 3.

### Data for regional Shapely value computation

To compute the contributions (Shapley values) of arterial, venous, and nonvascular regions to diagnosis of ocular and systemic diseases, two ocular diseases (HR, DR) and two systemic diseases (HF, MI) are involved in. We randomly selected 50 images of HR from the BRSET dataset. For DR, we randomly chose 30 images (comprising 7 of grade 1, 18 of grade 2, and 5 of grade 3) from the MESSIDOR-2, APTOS-2019, and IDRiD datasets. Additionally, 60 images of HF and 159 images of MI were randomly selected from the UKBB.

### Data for evaluating clinical consistency

To evaluate the consistency of interpretations among HRVRL, other foundational models, and expert annotations, we included two ocular diseases with well-established clinical standards: HR and DR. 30 images randomly selected of HR from the BRSET dataset. For DR, 30 images (comprising 7 of grade 1, 18 of grade 2, and 5 of grade 3) were randomly chosen from the MESSIDOR-2, APTOS-2019, and IDRiD datasets. Four ophthalmologists annotated the approximate regions where lesions appeared based on clinical diagnostic criteria^21,23^ using the LabelMe software^46^.

### Data for zero-shot lesion detection

In the zero-shot lesion detection task, we utilized the public IDRiD (India) dataset, which includes pixel-level annotation for microaneurysms, hemorrhages, hard exudates, and soft exudates of typical DR lesions^43^.

### Multi-level data augmentation strategy

We designed a novel data augmentation strategy to enhance resource efficiency with the prompt of vessel (**Extended Data Fig. 1**). Given a fundus image *X* and its binary mask *M*_*a*_and *M*_*v*_, six transformations were applied to create variations containing only vascular structures (arteries *X*_*a*_, veins *X*_*v*_, and both *X*_*av*_), non-vessel *X_nav_*, and non-vascular region with arteries *X_nv_*, or veins *X_na_*):

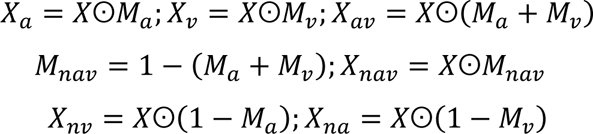

Where ʘ denotes element-wise bitwise multiplication. Furthermore, to capture vascular features across various spatial locations and scales, a multi-scale sampling strategy was applied: (1) a point was randomly selected on the retinal vascular skeleton, and (2) a large window *W*_2*P*_ of size 2*P* × 2*P* was centered at this point. Then, (3) a target patch *W*_*P*_ of *P* × *P* was selected within *W*_2*P*_. In our experiment setting, *P* was set to 96, 128, 256, 384 and 512, and the overlap between sample pairs of the same scale was restricted to 0.5 to prevent overfitting.

### HRVRL architecture and hierarchical pretraining

We proposed HRVRL, based on a novel hierarchical retinal vascular representation learning framework for analyzing fundus images (**Extended Data Fig. 3**). Inspired by the clinical observation that morphological changes in vessels, including arteries and veins, along with the appearance of lesions, may serve as early biomarkers for certain ocular and systemic diseases, we designed a hierarchical pretraining strategy for HRVRL. This strategy progressed from coarse-grained to fine-grained representations in two stages, capturing latent features that differentiate arteries, veins, and non-vascular regions. In the first stage, clinical knowledge was encoded into HRVRL by assigning vascular and non-vascular instances as positive and negative pairs for instance-level contrastive learning. In the second stage refined feature differentiation among arteries, veins, and non-vessels by pixel-level semantic segmentation.

Specifically, 267 fundus images with arterial and venous vessel labels were preprocessed to generate 158,842 instances. If an instance contained vascular information, it and its vascular-inclusive variations were defined as positive samples, its vascular-exclusive variations were defined as negative samples. If lacking vascular information, it was designated as a negative sample. During training, a normalized temperature-scaled cross-entropy loss function^47^ and AdamW optimizer^48^ were employed to update parameters. The learning rate was set to 4e-3, and batch size was set to 128. After 100 training epochs, 2,000 positive and negative sample pairs were randomly selected, and uniform manifold approximation and projection^49^ (UMAP) was used to evaluate the model’s ability to differentiate vascular from non-vascular features (**Extended Data Fig. 7A**).

In the pixel-level semantic segmentation stage, we incorporated our previous work RIP-AV^50^, which combines representative instance pretraining (RIP) and a context-aware network for artery/vein classification, to build the final HRVRL model (**Extended Data Fig. 3**). The original RIP, which lacked support for non-vascular instances, was modified by instance-level contrastive learning. In the training workflow, each patch and its surrounding context were input into a dual-stream encoder that processes these inputs to extract shallow and high-level feature maps. These feature maps were processed by the patch context fusion (PCF) module and the distance-aware (DA) module. The PCF module jointly applied self-attention and cross-attention to leverage relationships between local patches and their surrounding context, and DA module was designed with convolutional layers to enhance the perception of vessel edges. Finally, the decoder, using standard convolutional-batch normalization-ReLU (Conv-BN-ReLU) structures, produced a prediction map for arteries and veins (more details can be found in our previous work, RIP-AV). The generalization of HRVRL in distinguishing arterial, venous, and non-vascular regions was assessed and compared with current retinal arteries and vein classification tools using both internal and external data^51,52^ (**Extended Data Fig. 7B**).

### HRVRL adaption to downstream tasks

To accommodate specific downstream tasks, such as disease diagnosis and grading, the HRVRL decoder was removed, leaving only the encoder followed by a multilayer perceptron (MLP) for disease category prediction. Two adaptation approaches were employed: (1) extracting 384-dimensional embeddings from the frozen encoder and fitting an MLP classifier, and (2) fine-tuning all HRVRL layers. Before training, we removed or padded the black area of retinal images to form squares, then resized to 512 × 512 pixels with cubic interpolation. Subsequently, several augmentations were applied, including random flips, CutMix, grayscale, and normalization. A label smoothing cross-entropy loss function was used to regulate the output distribution. The AdamW optimizer was used with an initial learning rate set to 5e−4, a batch size of 16, and a total of 100 training epochs. To enhance the optimization process, we used a cosine learning rate scheduler that combines a warmup phase. More specifically, it increases the learning rate from 0 to 5e-4 over the first 10 epochs, followed by a cosine annealing schedule, which reduces the learning rate from 5e-4 to 1e-6 over the remaining epochs. All layers were updated using layer-wise adaptive learning rates and weight decay. The model checkpoint with the highest AUROC on the validation set was saved for evaluation.

### Regional to global interpretation of task-specific HRVRL

We proposed a novel hierarchical interpretation method that joint combines gradient-based saliency maps with Shapley values to enhance clinical explanation. The saliency map was generated using Grad-CAM^53^, which backpropagated class-specific gradients through the encoder’s final convolutional block, to highlight areas critical for model decisions. Shapley values quantified the importance of features by calculating the marginal contribution of individual features, considering all interactions with other features, and then averaging these contributions. This fair attribution method allocated the total prediction outcome to individual features, highlighting their contribution to the model’s decision-making process. Combining regional saliency maps with Shapley values could improve HRVRL’s explanation of fundus image analysis, enabling clinicians and researchers to visualize regional and global heatmaps and quantify the regional contribution to model predictions. The strategy is divided into four steps: (1) divide the fundus image into arterial region *r*_*a*_, venous region *r*_*v*_, and non-vascular region *r*_*n*_ based on the presence and type of vessels. (2) calculate prediction results for each region individually and in combination, then generate region-level saliency maps *s^region^*. (3) determine the marginal contribution of each region using the following formula. (4) construct the final explainable saliency map *s^fusion^* through a weight average and *min-max* normalization of the region saliency maps.

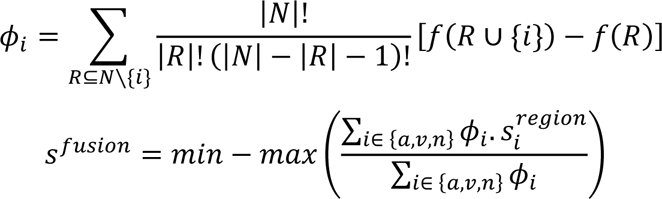

Where *ϕ*_*i*_ denotes the Shapley value for region *i*, *N* denotes the set containing all regions, i.e., *N* = { *r*_*a*_, *r*_*v*_, *r*_*n*_}. *R* is a subset of *N* that does not include *i*-th the region. |*N*| denotes the total number of regions, which is 3 in this case. *f*(*R*) is the prediction output for subset *R*. To evaluate the model’s interpretability and its concordance with clinicians, we assessed the similarity ratio between the number of regions highlighted in the heatmaps and the number of lesions annotated by experts.

### Detection of distinct types of lesions in a zero-shot manner

Evaluating model performance on zero-shot tasks is crucial for assessing robustness, adaptability, and generalization as it reflects the ability to effectively handle new tasks without prior training. To evaluate the zero-shot capabilities of HRVRL and other foundational models, we directly applied a diabetic retinopathy (DR) classification model to detect distinct lesion types. Specifically, we fine-tuned HRVRL on the MESSIDOR-2 dataset, a well-established DR severity grading dataset, and then applied it to detect various lesions on the IDRiD dataset without additional retraining. Interpretability heatmaps were generated and binarized with a threshold of 0.5. Accuracy was assessed by quantifying the spatial overlap between heatmaps and the various ground-truth lesion areas. Since Grad-CAM is typically used for CNN-based networks, the heatmaps of RETFound, DERETFound, and RETFound-Green are all calculated using RELPROP^54^, which is more suitable for Transformer-based networks.

### Ablation study of pretraining strategies

We conducted an ablation analysis by replacing the proposed pretraining strategy with three different initializations: random initialization, supervised learning pretraining on ImageNet-1k (SL-ImageNet), and instance-level contrastive learning of HRVRL (ICL-HRVRL) to generate variant pretrained models for comparison. For random initializations, we initialized all parameters of both convolutional and linear layers using the truncated normal distribution with a standard deviation of 0.02, which was the default setting in original ConvNeXt paper. For the SL-ImageNet initializations, we adopted the official supervised ConvNeXt-T weights based on the ImageNet-1K dataset^55^. For the ICL-HRVRL initializations, we directly employed the weights derived from the first stage of HRVRL, which were capable of distinguishing between vascular and non-vascular features. Then, we fine-tuned these pretrained models using the same protocol to adapt to downstream tasks and compare their performance.

### Computational resources assessment

We utilized only 267 public fundus images and, guided by the vascular structure, generated 158,842 instances featuring arteries, veins, and non-vessel regions for hierarchical pretraining. HRVRL, our model with a parameter size of approximately 49 MB (0.04 GB), was pretrained on a single NVIDIA A100 GPU with 80 GB of memory. The pretraining time comprised two parts: the first stage of instance contrastive pretraining took 15 hours, and the subsequent phase for further differentiation of arteries and veins took an additional 4.44 hours, totaling 19.44 hours, equivalent to 0.81 A100 days. To adapt HRVRL to specific classification tasks, we used a single NVIDIA RTX 4090 GPU (24 GB), which required approximately 30 minutes per 1,000 images. For inference speed evaluation, using a batch size of 1 and following the example script provided by RETFound, HRVRL processed each image in approximately 0.04 seconds.

### Performance Evaluation and statistical analysis

All diseases classification tasks were evaluated using widely adopted classification metrics including area under the receiver operating curve (AUROC) and the area under the precision-recall Curve (AUPR), accuracy, sensitivity, precision, specificity and F1 score. All quantitative results were represented in **Supplementary Table 3 and 4**. For binary disease diagnosis, we computed AUROC and AUPR in a binary setting. For multi-class ocular disease diagnosis and severity grading, we calculated AUROC and AUPR individually for each class and then averaged them using macro-averaging to derive the overall AUROC and AUPR scores. To ensure robustness of metrics and calculate p-values for fair comparisons between models, we bootstrapped the test dataset 1,000 times and then computed the AUROC and AUPR for each time, and reported the mean value and 95% confidence interval (CI). We used the 2.5% and 97.5% values to define the 95% CI. We employed two-sided t-tests to evaluate the statistical significance of the performance differences between the top two best-performing models. For non-normally distributed data, including lesions detection rate and type-specific lesion recall, we employed Mann–Whitney U-tests to evaluate the statistical significance of the performance differences between the top two best-performing models. The *P*-value below 0.05 indicates a significant difference.

## Data Availability

This study involved only 60 retinal images of MAGIC cohort in the pretraining stage and was approved by the Ethics Committees at the Eye Hospital of Wenzhou Medical University (2022-064-K-45-03) in accordance with the Declaration of Helsinki. All patients provided informed consent for the use of their retinal images.

## Data available

The pretraining datasets of AV-DRIVE, LES-AV, HRF and the downstream datasets can be accessed by referring to the original paper. Access to the MAGIC and UK Biobank (UKBB) pretraining datasets and their corresponding vascular labels requires contacting the authors. We aim to respond to such requests within two weeks.

## Code available

The code used to preprocess, pretrain, fine-tune and evaluate HRVRL will be available after being published.

## Author Contributions

The methodology was devised by W.D., Z.J.C., Y.Y., J.S., and J.Q. The foundation model was built by W.D., Z.J.C., Y.Y., J.S., Y.C. and J.F. The experiments were performed by W.D., Z.J.C., Y.Y., Y.C., J.F., Q.B., C.X., H.W. and H.D. The results mentioned in the paper were analyzed by W.D., Z.J.C, Y.Y., J.S., Y.Y. and T.H. The annotation of retinal vessels and lesions were contributed by H.Y., R.Z., R.Z. and C.Y. The MAGIC data and computing facilities were provided by J.Y., H.W., L.X. and X.Y. The manuscript was written by W.D., Z.J.C., Y.Y., J.S., and J.Q. with contributions from all other authors.

## Acknowledgements

We thank Prof. Fan Lyu and Kang Zhang for their constructive comments regarding this manuscript. This work was supported by the National Natural Science Foundation of China (81830027, U20A20364) and the Zhejiang Provincial Key Research and Development Program Grant (2021C03102) to J.S.; the National Natural Science Foundation of China (82172882) to J.S. We also thank the authors of RETFound, DERETFound, as well as RETFound-Green, for their contribution to the field and particularly for making their models openly available which enables the comparisons in this work. We acknowledge the UK Biobank for their contribution of data and resources that supported this research. We further thank the researchers that made the datasets used in this work available and the individuals who contributed their data to biomedical research.

## Competing interests

The authors declare no competing interest.

---

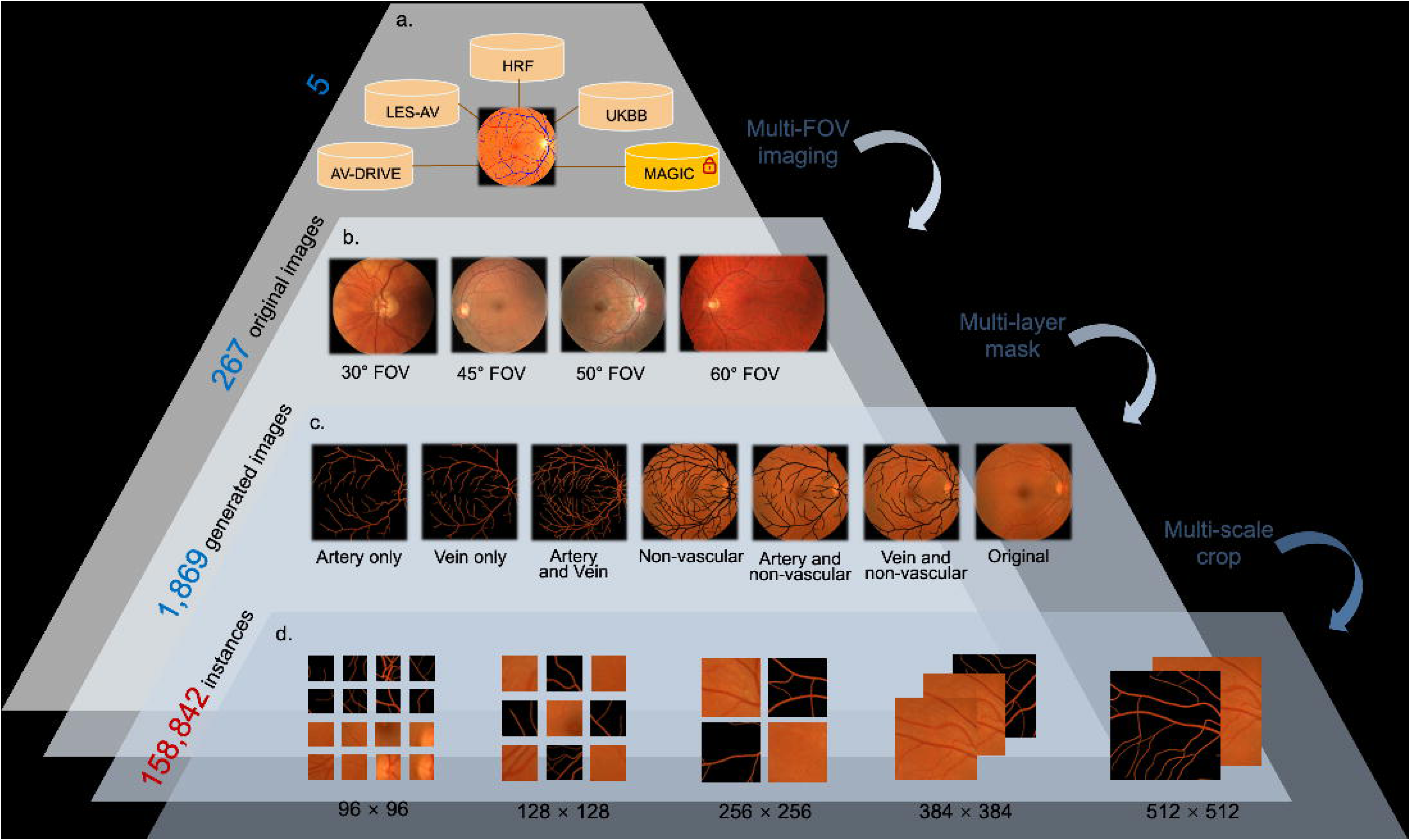

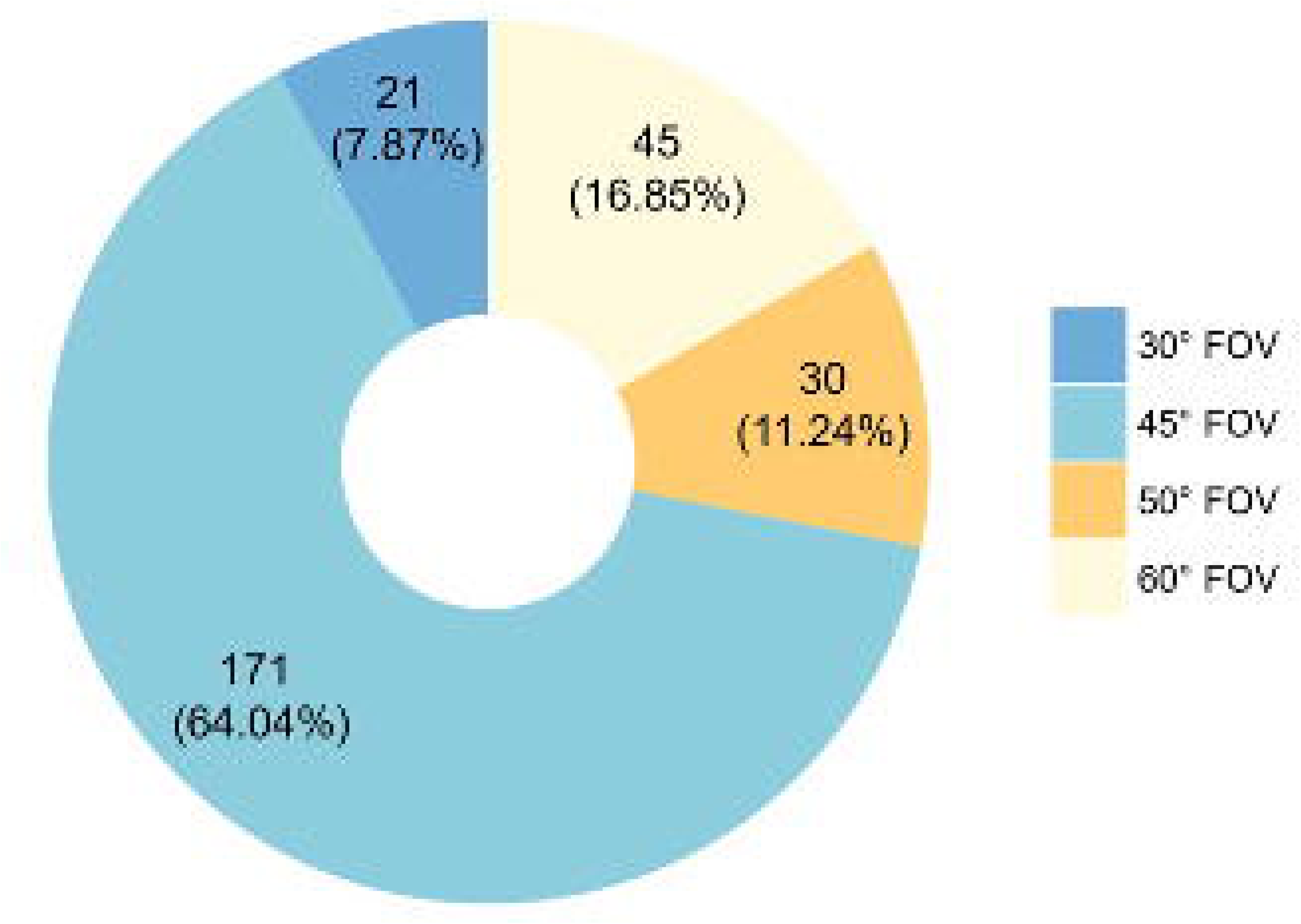

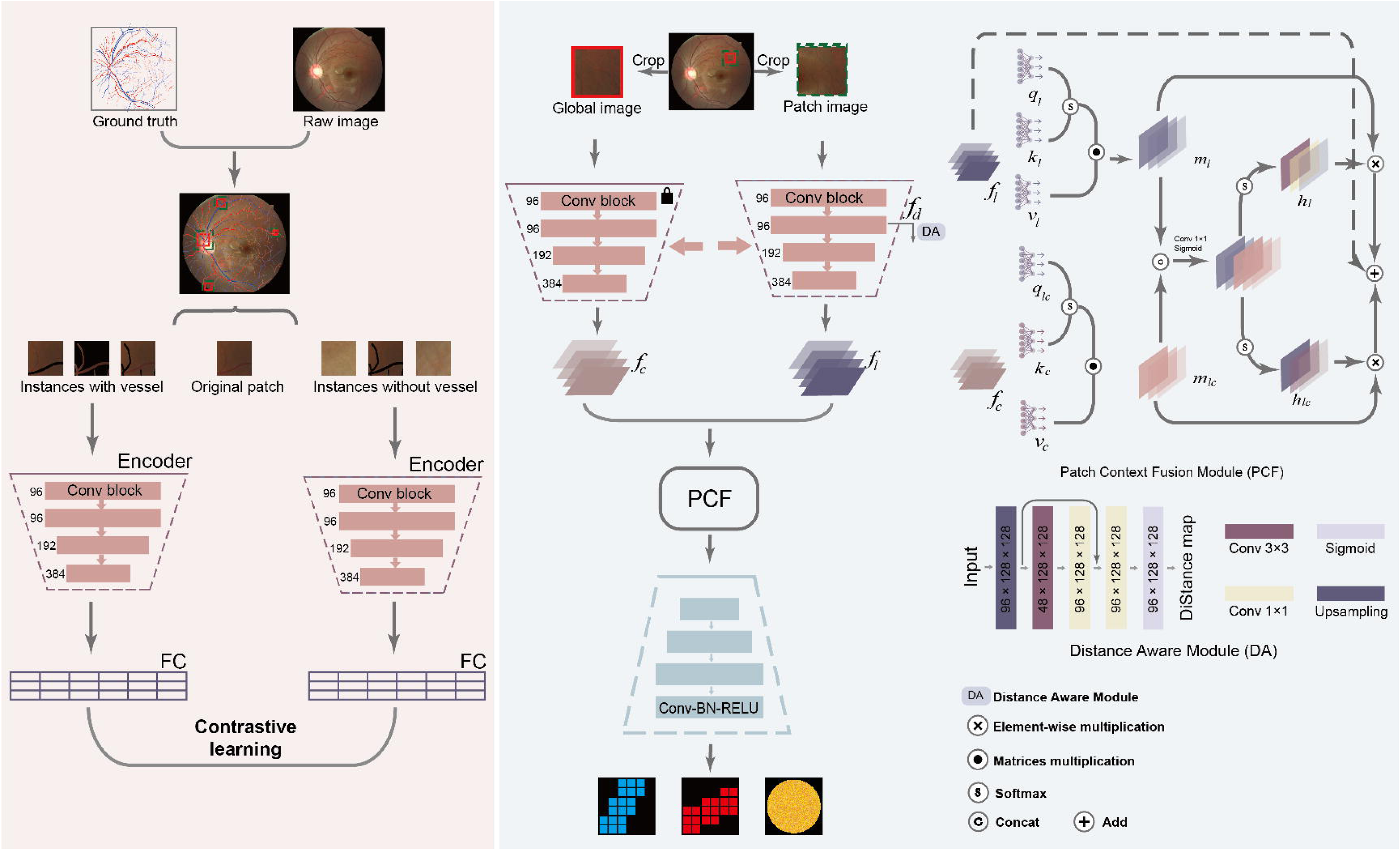

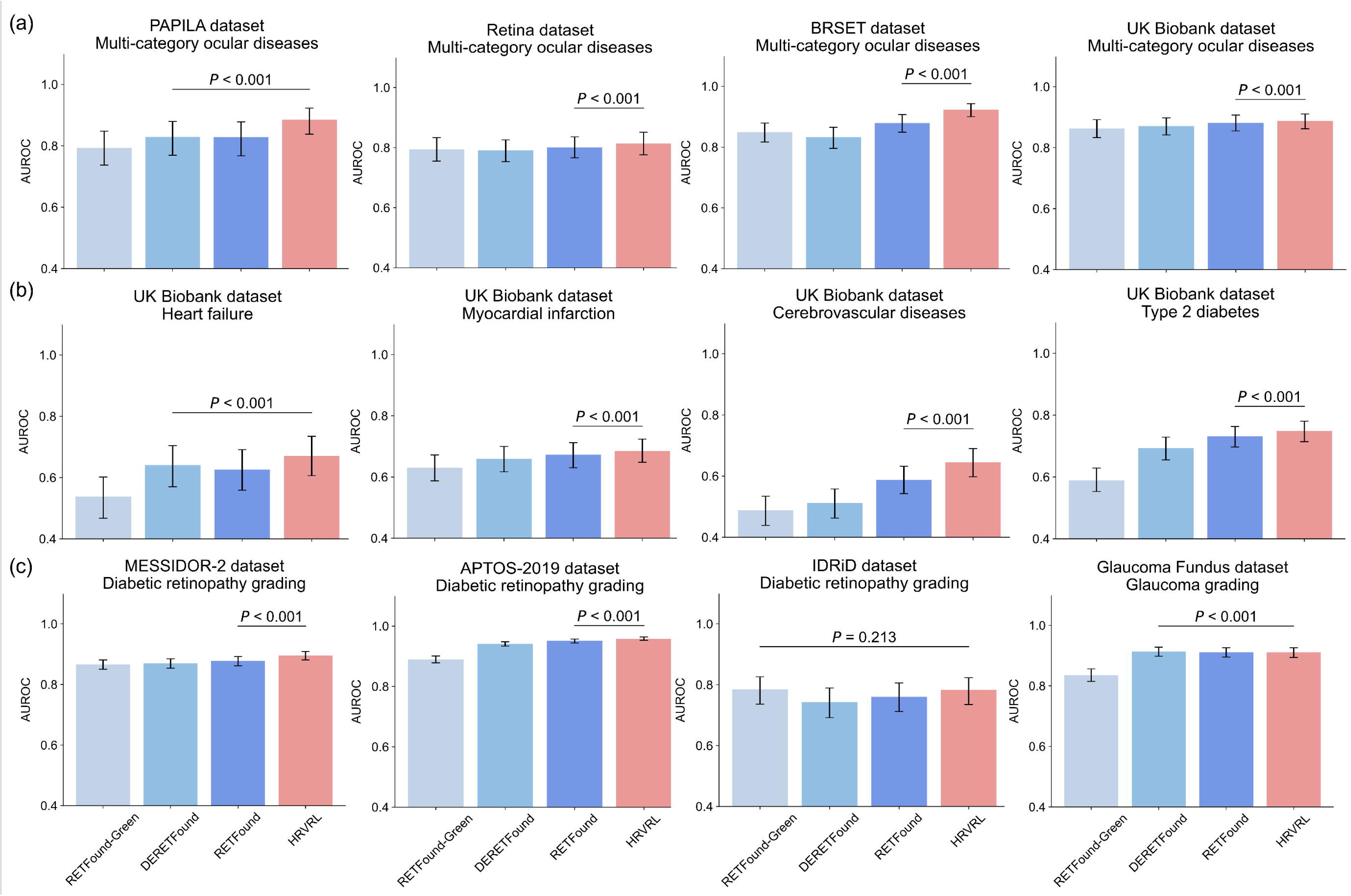

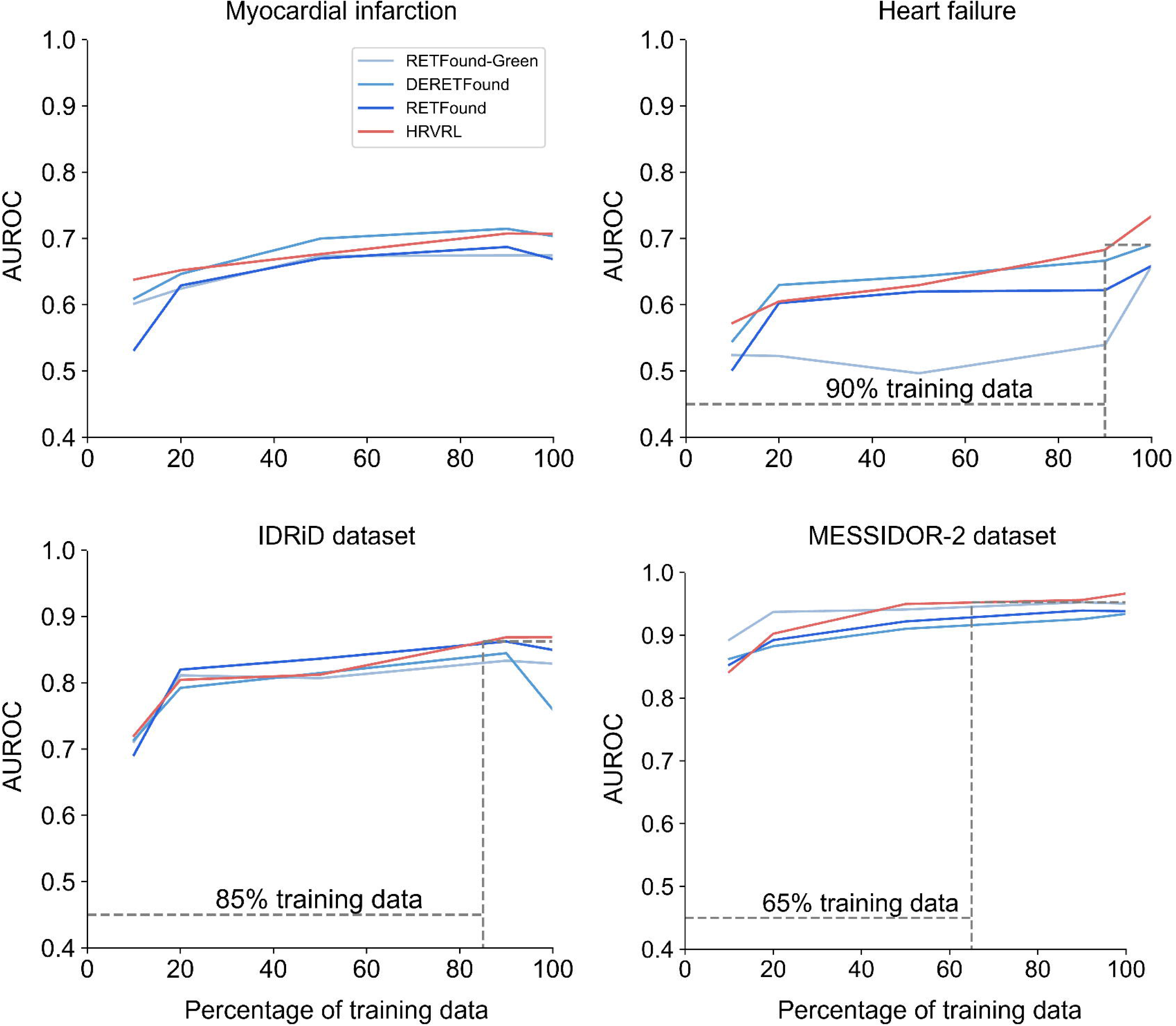

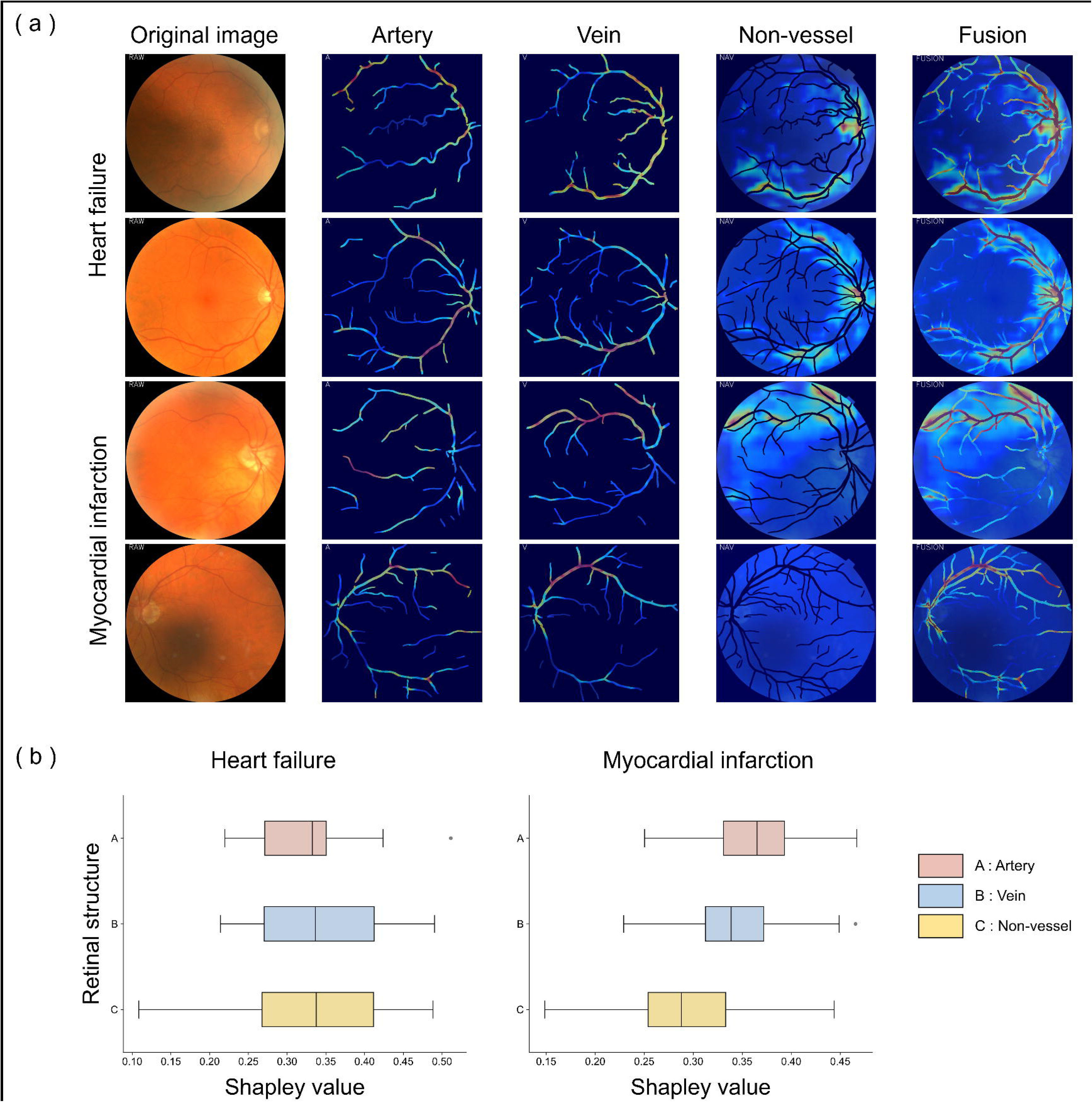

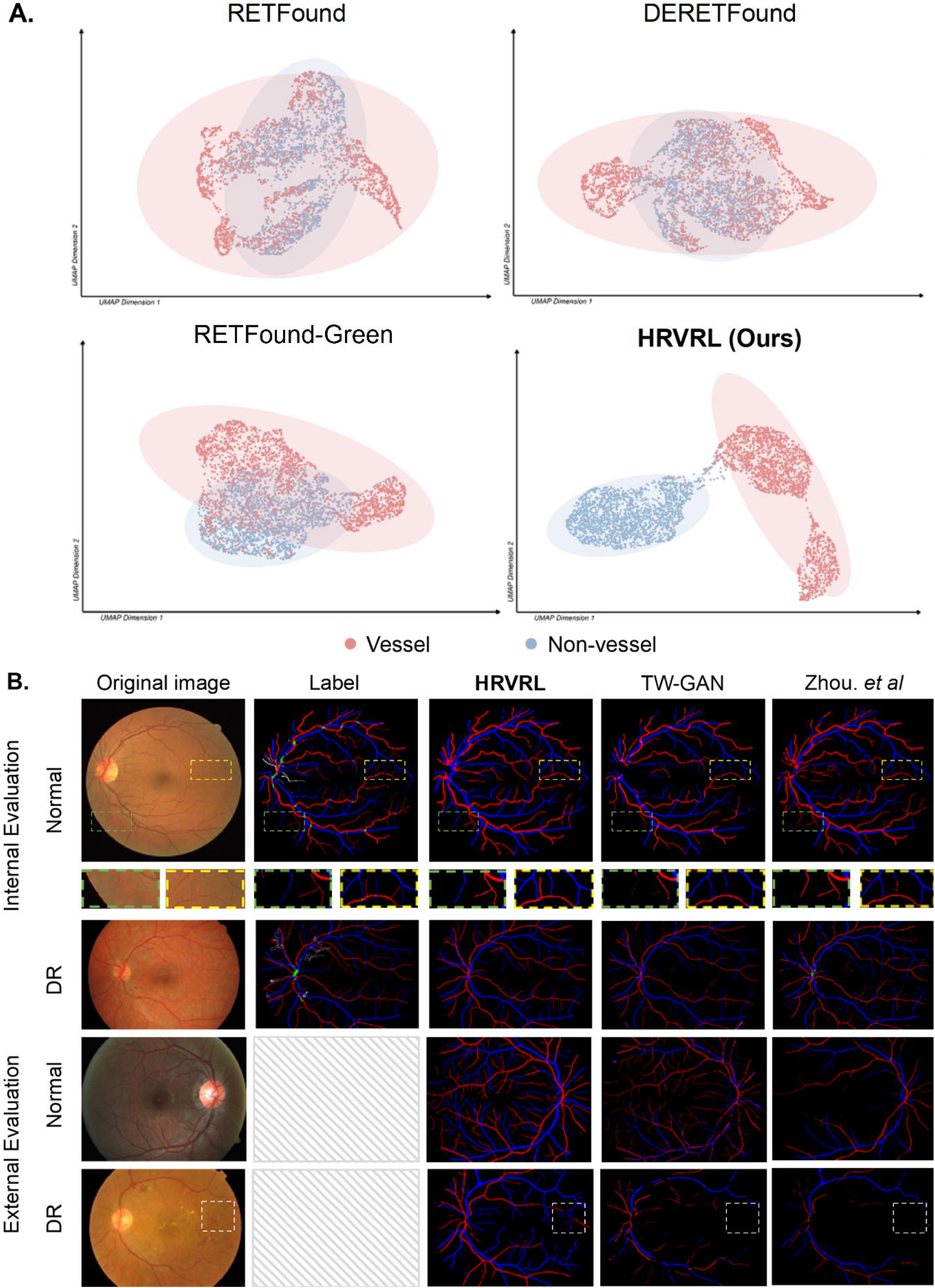

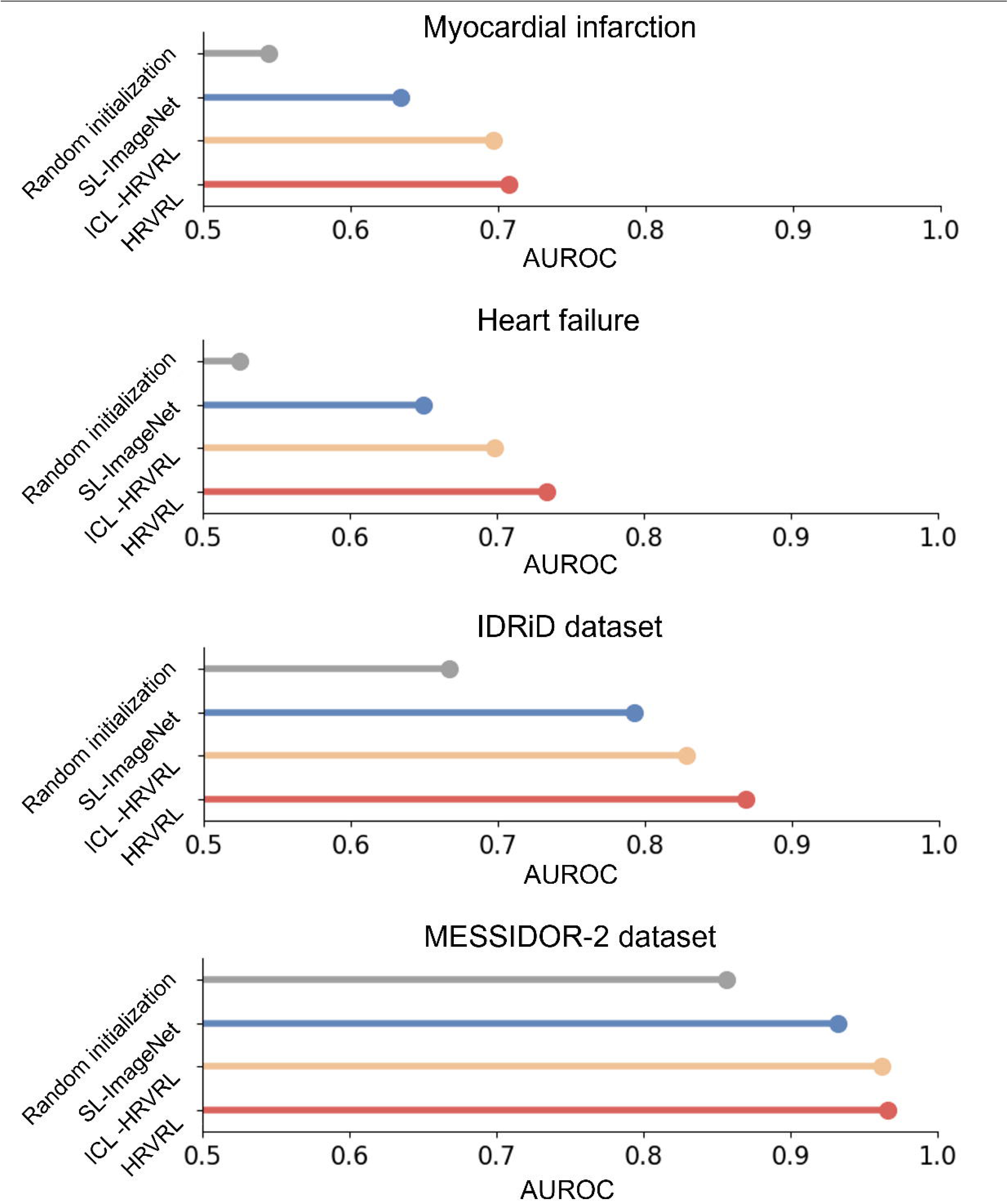

## Notes

### Competing Interest Statement

The authors have declared no competing interest.

